# Laboratory biomarkers associated with COVID-19 severity and management

**DOI:** 10.1101/2020.08.18.20168807

**Authors:** S Keddie, O Ziff, MKL Chou, RL Taylor, A Heslegrave, E Garr, N Lakdawala, A Church, D Ludwig, J Manson, M Scully, E Nastouli, MD Chapman, M Hart, MP Lunn

## Abstract

The heterogeneous disease course of COVID-19 is unpredictable, ranging from mild self-limiting symptoms to cytokine storms, acute respiratory distress syndrome (ARDS), multi-organ failure and death. Identification of high-risk cases will enable appropriate intervention and escalation. This study investigates the routine laboratory tests and cytokines implicated in COVID-19 for their potential application as biomarkers of disease severity, respiratory failure and need of higher-level care.

From analysis of 203 samples, CRP, IL-6, IL-10 and LDH were most strongly correlated with the WHO ordinal scale of illness severity, the fraction of inspired oxygen delivery, radiological evidence of ARDS and level of respiratory support (p≤0.001). IL-6 levels of ≥3.27pg/ml provide a sensitivity of 0.87 and specificity of 0.64 for a requirement of ventilation, and a CRP of ≥37mg/L of 0.91 and 0.66.

Reliable stratification of high-risk cases has significant implications on patient triage, resource management and potentially the initiation of novel therapies in severe patients.

## Introduction

International efforts to ‘flatten the curve’ of COVID-19 through social isolation restrictions have allowed most health systems to cope with huge demands on healthcare facilities, in particular providing time to acquire ventilators and intensive care beds. As governments loosen lockdown processes and with the unknown influences of seasonal variation and acquired immunity, front line services need to prepare for a second wave of infection. Lessons learnt regarding COVID-19 disease biology, transmission, risk factors, complications and treatments will be used to adapt and improve clinical services and improve outcomes.

The heterogeneous disease course of COVID-19 is unpredictable with most patients experiencing mild self-limiting symptoms. However up to 30% require hospitalisation, and 17% of such require intensive care support for acute respiratory distress syndrome (ARDS), hyperinflammation and multiorgan failure.^1–3^ A cytokine storm in patients with severe disease was identified in the early reports of Wuhan patients and is intrinsic to disease pathology. In this cohort, elevated plasma IL-2, IL-7, IL-10, GCSF, IP10, MCP1, MIP1A and TNF-α levels in ICU patients were identified. ^4^ Some subsequent studies have implicated IL-6 as a valuable predictor of adverse clinical outcome and a potential therapeutic target. ^5,6^ One or more clinical and wet biomarkers may enable early identification of high-risk cases, assisting disease stratification and effective use of limited specialist resources.

This study comprehensively explored clinical disease features and routine laboratory tests against specialist cytokine biomarkers associated with COVID-19 disease and its complications, to address their association with disease severity, respiratory intervention and outcome.

## Methods

One hundred consecutive hospital in-patients with COVID-19 infection, whose sera were sent for cytokine testing, were investigated as part of standard care and routine practice at University College London Hospital NHS Trust. All cases were confirmed as COVID-19 by PCR analysis (86%) or deemed highly likely based on internationally recognised diagnostic criteria including recent exposure, clinical, radiological and laboratory features. ^7^ Laboratory biomarkers including cytokines were collated with baseline demographics, risk factors, disease features, treatments and outcomes. Level of clinical care (ward based, high-dependency or intensive treatment), oxygen requirements and vital signs were associated with the biomarkers at the time of sample collection, at peak illness and on most recent assessment. Serial samples were analysed to establish whether biomarkers correlated with, or could predict disease course. Severity of COVID-19 illness was determined utilising the WHO COVID-19 ordinal severity scale, with a score of 1 defined by no limitation of activities, to 7 requiring ventilation and additional organ support. ^8^

Bloods for cytokine analysis were centrifuged within 4 hours of collection, separated and sera frozen at −80°C for up to 24 hours before being analysed. Immunoassays were performed according to manufacturer’s instructions and reads were fully automated. Routine laboratory data from automated analysers including CRP, D-dimer, LDH, lymphocyte count, ferritin, fibrinogen and platelets were collated with the clinical and cytokine data.

A four-PLEX cytokine ELISA-based electrochemiluminescent immunoassay (Meso Scale Discovery, Rockville, Maryland) was introduced on the 17^th^ April 2020 for clinical use in COVID-19 cases. The immunoassay measures IL-6, IL-1β, IL-10 and TNF-α to levels μ1pg/ml, with the choice of the four analytes determined by preliminary published reports, known disease pathogenesis and treatment considerations. The acquired data were part of assay validation and verification to provide normal and disease ranges and clinical interpretation.

### Statistical analysis

Chi-square was used for the comparison of categorical variables. Biomarker levels comparing dichotomous variables were evaluated using Welch’s unpaired t-test. Comparison of multiple group outcomes was performed using one-way ANOVA. Biomarker and continuous clinical variables were correlated using Pearson correlation coefficient. The predictive values of biomarkers were calculated by identifying Youden’s index on receiver operator characteristic curves. P-value ≤0.05 was considered statistically significant. All statistical analyses were made using R statistical package 3.6.2.

## Results

Samples from 100 patients were collected from the 6^th^ April to the 18^th^ May from patients hospitalised at UCLH sites. Eighty-six patients were COVID-19 PCR positive and 14 deemed highly likely based on diagnostic criteria. ^7^ Seventy-four cases were male, with a median age of 59 years (20-92). One hundred and forty samples were collected from ward environments versus 63 on intensive care. The WHO COVID severity score ranged from 3 (hospitalised, no oxygen therapy) to 7 (severe disease, ventilation and additional organ support) with a median of 6 (severe disease, intubated and ventilated). Background characteristics associated with more severe infection and intervention included older age (p≤0.001), diabetes (p≤0.05) and hypertension (p≤0.05). Clinical features and grouped biomarkers are displayed in Table 1

**Table 1:** Demographic characteristics, features of COVID-19 infection and biomarkers stratified against the WHO COVID-19 ordinal severity scale.

| | All patients | WHO 3<br>Hospitalised | WHO 4 & 5<br>Oxygen ± NIV | WHO 6 & 7<br>Ventilated | P-value |
| --- | --- | --- | --- | --- | --- |
| Number of patients | 100 | 24 | 23 | 53 |  |
| **Demographics** |  |  |  |  |  |
| Age (years) | 59 (20-92) | 59 (20-90) | 64 (24-92) | 57 (33-71) | $\leq 0.0001$ |
| Male | 74 (75%) | 17 (71%) | 16 (70%) | 41 (77%) | 0.715 |
| **Comorbidities** |  |  |  |  |  |
| Diabetes | 25 | 9 | 1 | 14 | $\leq 0.005$ |
| Hypertension | 37 | 9 | 8 | 19 | **0.005** |
| Cardiovascular disease | 12 | 4 | 5 | 2 | 0.53 |
| Cerebrovascular disease | 8 | 4 | 3 | 0 | 0.16 |
| Malignancy | 10 | 3 | 3 | 2 | 0.88 |
| Current smoker | 13 | 4 | 3 | 6 | 0.632 |
| **Clinical features** |  |  |  |  |  |
| Fever on admission | 84 | 19 | 18 | 47 | $\leq 0.0001$ |
| Rigors | 21 | 2 | 7 | 12 | **0.03** |
| Cough | 77 | 17 | 17 | 43 | $\leq 0.0001$ |
| Breathlessness | 73 | 10 | 20 | 43 | $\leq 0.0001$ |
| Sputum | 15 | 1 | 7 | 7 | 0.091 |
| Fatigue | 32 | 5 | 7 | 20 | $\leq 0.01$ |
| Myalgia | 16 | 5 | 2 | 9 | 0.099 |
| Anosmia | 2 | 1 | 1 | 0 | 0.61 |
| Sore throat | 10 | 3 | 1 | 6 | 0.15 |
| Headache | 12 | 6 | 2 | 4 | 0.37 |
| Diarrhoea | 15 | 5 | 1 | 9 | $\leq 0.05$ |
| Nausea | 15 | 4 | 2 | 9 | 0.074 |
| Vomiting | 9 | 3 | 0 | 6 | $\leq 0.050$ |
| Normal chest x-ray | 18 | 16 | 2 | 0 | $\leq 0.0001$ |
| Focal consolidation | 8 | 4 | 4 | 0 | 0.13 |
| Bilateral consolidation | 74 | 4 | 17 | 53 | $\leq 0.0001$ |
| **Treatment** |  |  |  |  |  |
| Dialysis | 17 | 0 | 1 | 16 | $\leq 0.0001$ |
| Antibiotics | 75 | 7 | 21 | 47 | $\leq 0.0001$ |
| Antivirals | 7 | 1 | 2 | 4 | 0.37 |
| Steroids | 28 | 3 | 4 | 21 | $\leq 0.0001$ |
| **Biomarkers** |  |  |  |  |  |
| IL-1β pg/ml | 0.4 | 0.7 | 0.2 | 0.4 | $\leq 0.05$ |
| IL-6 pg/ml | 50.8 | 5.9 | 16.1 | 85.4 | $\leq 0.0001$ |
| TNFα pg/ml | 5.2 | 4.1 | 4.4 | 6.1 | 0.10 |
| IL-10 pg/ml | 3.4 | 1.5 | 3.2 | 4.4 | $\leq 0.0001$ |
| Lymphocyte $\times 10^9/L$ | 1.4 | 1.4 | 1.4 | 1.4 | 0.22 |
| CRP mg/L | 102.7 | 49.4 | 73.3 | 135.7 | $\leq 0.0001$ |
| D-dimer ug/L | 7131 | 4007 | 6490 | 8732 | $\leq 0.0001$ |
| Ferritin ug/L | 1470 | 945 | 2129 | 1436 | $\leq 0.001$ |
| LDH IU/L | 406.5 | 311.0 | 378.4 | 451.7 | $\leq 0.001$ |
| Fibrinogen g/L | 17.6 | 4.2 | 5.0 | 26.0 | $\leq 0.001$ |
| Platelets $\times 10^9/L$ | 295.4 | 274.9 | 274.5 | 314.0 | 0.24 |

IL-1β was the only biomarker which significantly differentiated by gender, with higher levels in males (p≤0.01) who have a higher mortality.^9^ Biomarkers of COVID-19 severity did not differ by number of comorbidities or smoking status.

All the biomarkers in figure 1 were analysed for their correlations with WHO COVID-19 severity scores and arranged into a heat map (See Figure 1). Biomarkers that significantly correlated with the WHO severity score included CRP [*R=* 0.53, p=2.6e^−12^), IL-6 (*R* 0.49, p = 1.3e^−13^), LDH (*R* 38, p < 2.1e~^−5^) and IL-10 *(R* 0.35, p = 2.5e^−7^). IL-6 and CRP were most significantly indicative of the level of respiratory support (Figure 2a and 2b) with IL-6 being marginally better at differentiating respiratory requirement from none. I1-6 and CRP were both closely associated with increased fraction of inspired oxygen delivery (FiO2) requirements (R=0.54 and 0.58 respectively) and radiological evidence of ARDS (IL-6≤0.00001, CRP ≤0.005). A cut-off of 3.27pg/ml for IL-6 gives a sensitivity 0.87 and specificity 0.64 for requirement of Intubation. Similarly, a 37mg/l cut-off for CRP has a sensitivity of 0.91 and specificity of 0.66. TNF-α also correlated with WHO severity (p≤0.05) and level of respiratory support (p≤0.01) but to a lesser degree than IL-6 and CRP. A requirement for renal dialysis was determined by higher levels of TNF-α (mean 5.87 vs 4.7pg/ml, p<0.01), CRP (162 vs 82mg/L, p<0.01), IL-6 (1.1 vs 0.6pg/ml, p<0.01) and IL-10 (3.9 vs 2.44pg/ml, p<0.05). IL-1β did not correlate with disease severity measures, presence of ARDS or level of respiratory support.

**Figure 1:**
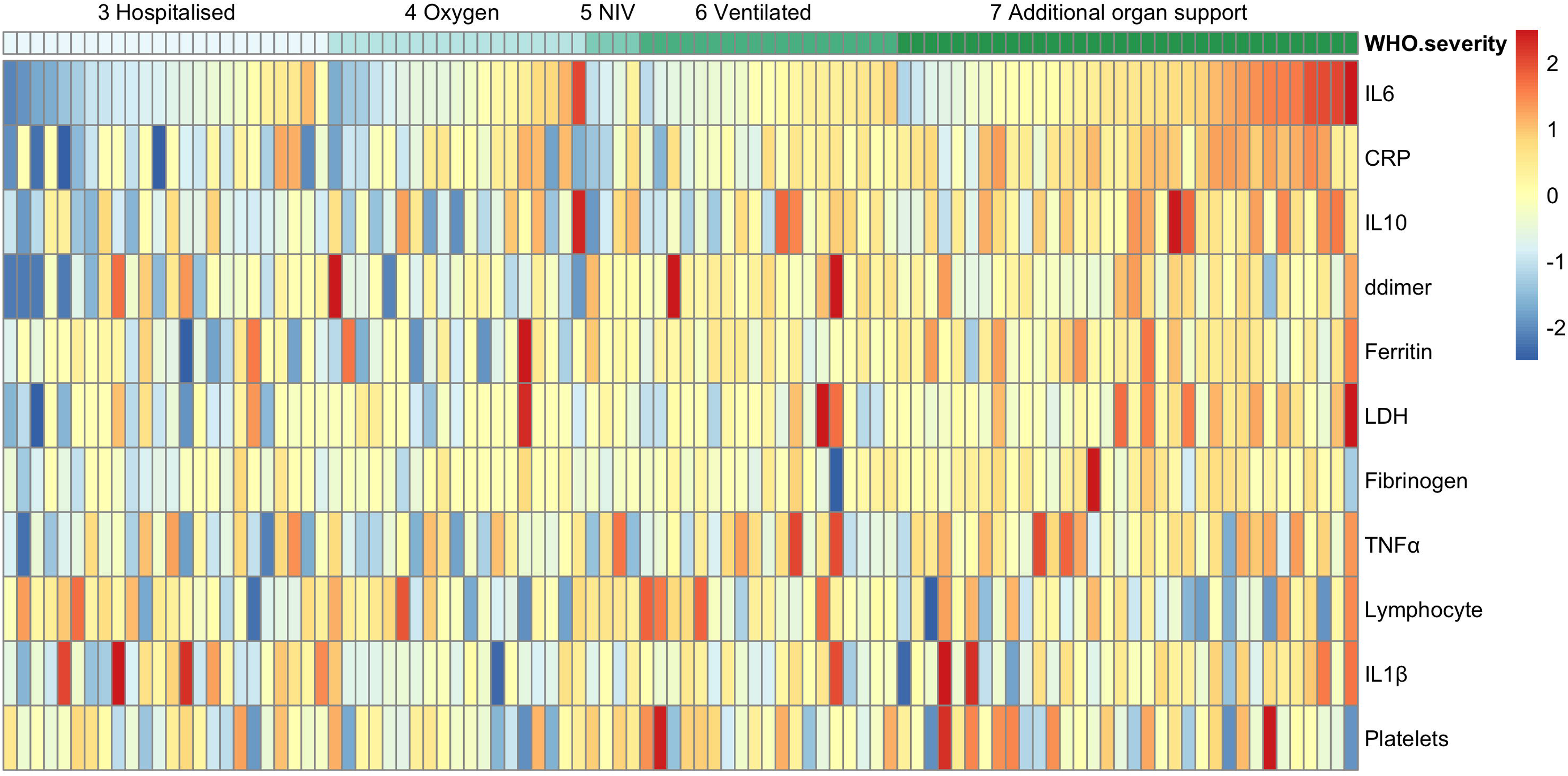
Heatmap of biochemical markers and their association with COVID-19 illness severity. Heatmap shows biochemical signatures for each patient, arranged by WHO COVID-19 severity score. Visualisation was performed using the *pheatmap* R package. Biomarkers values are log10 transformed, centred and scaled. No patients had WHO severity scores 0,1 or 2 as these are non-hospitalised patients. A score of 3 is hospitalised patients requiring no oxygen therapy; a score of 4 requires oxygen therapy; 5 requires non-invasive ventilation (NIV); of 6 requires intubation and mechanical ventilation; and 7 requires ventilation and additional organ support including vasopressors, renal replacement therapy and ECMO. A score of 8 is death. Higher levels of CRP, IL-6, IL-10 LDH and TNF-α are associated with higher WHO COVID-19 severity scores.

**Figure 2:**
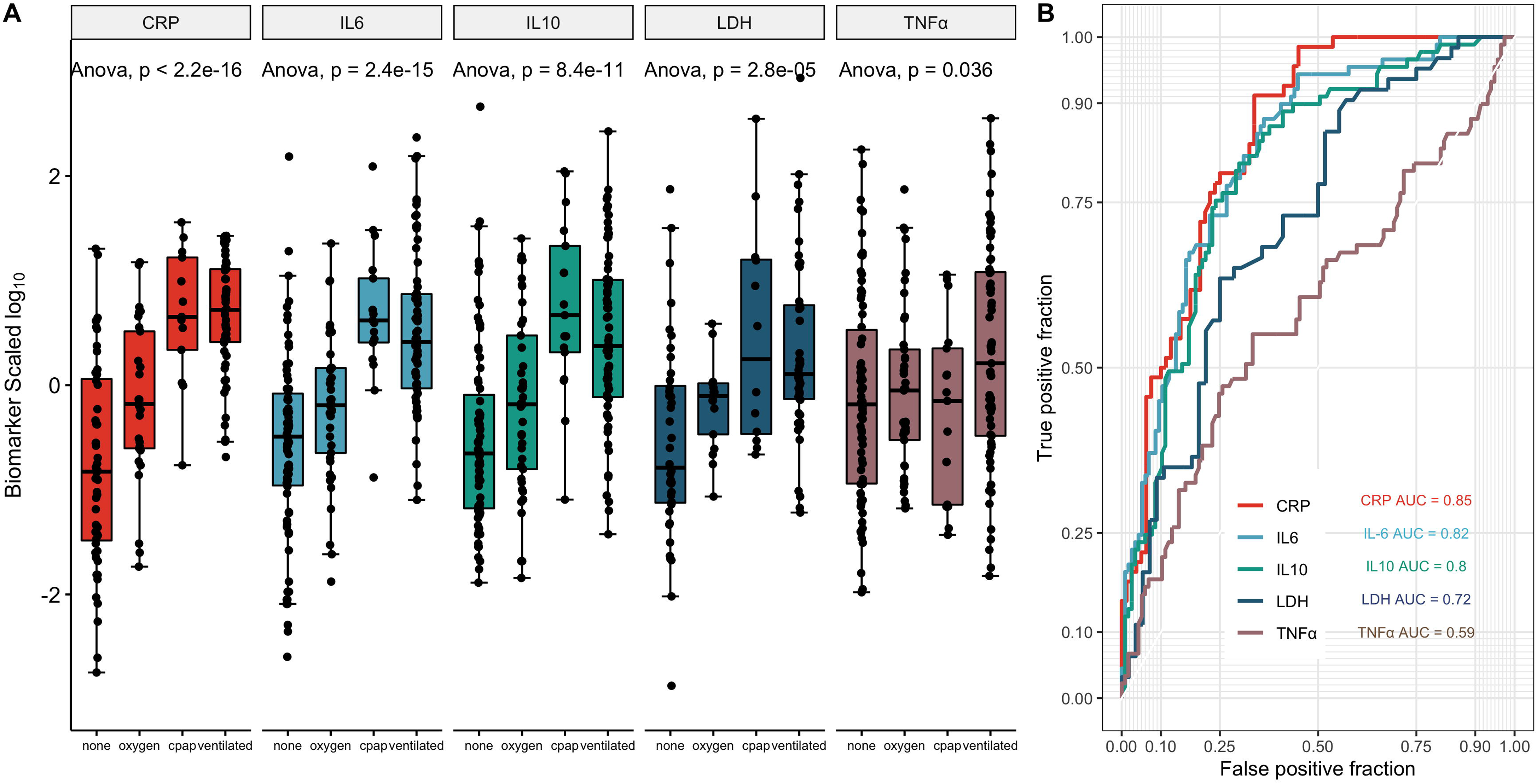
CRP, IL-6, IL-10, LDH and TNF-α levels predict the requirement for mechanical ventilation and intensive care. A) Split by four categories of respiratory support (none; supplemental oxygen; continuous positive airway pressure (CPAP); and mechanical ventilation. One way ANOVA shows these four categories are significantly different for CRP, IL-6, IL-10, LDH and TNF-α. B) Receiver operator characteristic curves of CRP, IL-6, IL-10, LDH and TNF-*α* demonstrating the area under the curve (AUC) for predicting the requirement for intensive care.

The strongest correlation between two biomarkers was unsurprisingly CRP and IL-6 (R=0.65 p≤0.00001) as CRP production is driven directly by IL-6, however all 4 cytokines analysed correlated significantly with one another (data not shown). D-dimer, ferritin and lymphocytes (inversely) correlated with all cytokine levels, indicative of severe COVID-19 resulting in a frank proinflammatory cytokine storm.

Longitudinal serial serum samples were also collected in 56 cases. Although not statistically significant, IL-6 and CRP levels trended downwards in patients located in ward environments compared to those in ITU in which levels remained high, and increased in those who subsequently died (data not shown). Serial levels of other biomarkers did not significantly correlate with clinical indicators of improvement i.e. as patients’ oxygenation improved or were stepped down from ITU.

## Discussion

With 15.2 million COVID-19 confirmed worldwide cases and over 620,000 deaths by the 7^th^ July 2020,^10^ understanding of COVID-19 disease pathogenesis, stratification and management has become a global priority. The identification of patients’ ventilatory requirements has been paramount to healthcare structuring and resource management Clinical assessment of COVID-19 cases remains key to decision making, but laboratory biomarkers enable finesse of this Information assisting decision making and provide data of underlying biological processes.

This study has comprehensively analysed the range of COVID-19 biomarkers for their usefulness in COVID-19 disease stratification. We have demonstrated that IL-6, CRP, IL-10, LDH and TNF-α are indicative of different aspects of COVID-19 severity; oxygen requirement, presence of ARDS and requirements of intensive care support including dialysis and ventilation. IL-6 is a potent proinflammatory cytokine which stimulates CRP and has been identified as a driver of the COVID-19 cytokine storm associated with poor outcome. ^5,6,11–13^ Herold *et al* have recently found an IL-6 cut-off value of 80pg/ml in their assay is highly indicative of respiratory failure, misclassifying only one patient not requiring intubation. ^5^ Such a clear definition has not been replicated in this study, which could be due to differences in assays or sample populations. Regardless, both studies demonstrate the significant utility of IL-6 as a key biomarker of disease in COVID-19. This supports the rationale behind the trial use of anti-IL-6 monoclonals as one of the therapies for COVID-19.^14,15^ Pre-selecting patients with the worst prognosis and most severe disease on the basis of highly elevated IL-6 may justify anti-interleukin immune therapy and avoid unnecessary complications by avoiding treatment in others.

IL-10 is a macrophage activated immunoregulatory cytokine which inhibits the expression of Th1 cytokines and enhances B cell stimulation. Elevated levels are therefore likely in response to the proinflammatory cytokine milieu, inherent to severe disease and reflect a functioning immune system.^16^ The binding of COVID-19 to the Toll Like Receptor (TLR) causes release of pro-IL-β which is cleaved by caspase-1, followed by inflammasome activation and production of active IL-β which is a mediator of lung inflammation, fever and fibrosis. IL-β has been shown to be raised in COVID-19 infection compared to healthy controls by some. ^4^ This study however does not provide any indication that IL-β levels reflect severity of disease.^4,17^ Despite this, initial trials of the IL-β antagonist Anakinra appear to reduce the need for mechanical ventilation and mortality in severe cases.^18^ However no studies have studied the influence of the drug on pre- and post-treatment levels of IL-β, or stratified entry to Anakinra on IL-β levels. Our study would indicate that non-IL-β biomarkers might be of more practical utility.

The association of TNF-α, CRP and IL-10 with dialysis requirement without any association with IL-6 possibly supports a role for sepsis driving renal failure. In post-mortem studies microthrombi are seen in both the kidney and alveolar blood vessels, but the primary insult on those two tissues may be different.

C-reactive protein is unsurprisingly very strongly correlated with levels of IL-6, as IL-6 drives the production of CRP primarily in the liver. CRP may potentially suffice in isolation to predict high risk cases requiring more aggressive intervention, negating the requirement of more specialist cytokine tests. Similarly, LDH, released from multiple tissues on cell death and activated through T-cell proliferation may be a practical and easily measured surrogate biomarker although its utility here is lower than the CRP. Levels of ferritin were raised across the cohort likely reflecting heightened acute phase response to infection as opposed to supra-high levels indicative of sHLH. Although select routine laboratory markers correlate with disease severity, cytokine analysis provides additional predictive support for prognosis and likely interventions in the COVID-19 cytokine storm as well as reassurance that when low, little escalation of intervention may be required. Note that a limitation of the study is that 16 cases were deemed not appropriate for intensive care, therefore escalation of respiratory support would be precluded regardless of clinical severity. We conducted a sensitivity analysis and inclusion/exclusion of these cases had no effect on biomarker correlations with severity.

Biomarker analysis of CRP, LDH and the cytokines IL-6, IL-10 and TNFα, alongside thorough clinical assessment of COVID-19 patients, enables more accurate stratification of high from low risk cases and the need for intensive care support. Such stratification enhances management not only for individual patients but has significant implications on resource management, hospital flow and staffing, and potentially the initiation of novel therapies in the most severe patients.

## Data Availability

Data is available upon the request of the corresponding author

